# Comparison between one and two dose SARS-CoV-2 vaccine prioritisation for a fixed number of vaccine doses

**DOI:** 10.1101/2021.03.15.21253542

**Authors:** Edward M. Hill, Matt J. Keeling

## Abstract

The swift development of SARS-CoV-2 vaccines has been met with worldwide commendation. However, in the context of an ongoing pandemic there is an interplay between infection and vaccination. Whilst infection can grow exponentially, vaccination rates are generally limited by supply and logistics. With the first SARS-CoV-2 vaccines receiving medical approval requiring two doses, there has been scrutiny on the spacing between doses; an elongated period between doses allows more of the population to receive a first vaccine dose in the short-term generating wide-spread partial immunity. Focusing on data from England, we investigated prioritisation of a one dose or two dose vaccination schedule given a fixed number of vaccine doses and with respect to a measure of maximising averted deaths. We optimised outcomes for two different estimates of population size and relative risk of mortality for at-risk groups within the Phase 1 vaccine priority order. Vaccines offering relatively high protection from the first dose favour strategies that prioritise giving more people one dose, although with increasing vaccine supply eventually those eligible and accepting vaccination will receive two doses. Whilst optimal dose timing can substantially reduce the overall mortality risk, there needs to be careful consideration of the logistics of vaccine delivery.

## Introduction

Vaccination has been seen as a key tool in the fight against SARS-CoV-2, although deployment provides multiple unique challenges that are not encountered by other vaccination programmes. In short, there is a race between infection and vaccination, with vaccination rates currently limited by supply and logistics, whereas infection can grow exponentially.

The vaccines developed represent a major technological achievement and have been shown to generate significant immune responses, as well as offering considerable protection against disease [1–5]. Field data from Israel and the UK suggested that protection against severe disease (hospitalisation or death) may be even greater [6, 7].

The data and science surrounding the SARS-Cov-2 infection is fast moving, so much so that publications can rarely keep pace. This paper was originally written in January 2021, to address contemporary public health questions. As such, this manuscript is largely a record of the state of our modelling at that time, although we interpret the results in terms of the latest data and policy questions.

At the original time of writing, in the UK the two vaccines currently deployed as part of the vaccination programme were the Pfizer/BioNTech and Oxford/AstraZeneca vaccines. The mRNA Pfizer/BioNTech vaccine was approved by the Medicines and Healthcare products Regulatory Agency (MHRA) on 2^nd^ December 2020 [8]. The Oxford/AstraZeneca vaccine, a chimpanzee adenoviral vectored vaccine, has been the main component of the UK vaccination program since it received approval for use by the MHRA on 30th December 2020 [9]. Both require two doses to be administered to maximise efficacy and longevity of immunity (with the duration of vaccine-derived immunity still uncertain).

A key question, given the urgency to achieve high levels of protection in the population, is the appropriate interval between first and second doses – often conceptualised as prioritisation of first or second doses. A longer interval allows more people to be given partial protection from one dose over relatively short-time scales, whereas a shorter interval will provide greater (although not complete) protection to the most vulnerable. In deciding between these two options, a number of factors need to be considered: the relatively high efficacy of the first dose from 3-12 weeks after vaccination [10]; the high levels of SARS-CoV-2 prevalence [11], COVID-19 morbidity [12] and COVID-19 mortality [13] since the start of the vaccine programme; the evidence that the Oxford/AstraZeneca vaccine provides greater second dose efficacy with a spacing of 12 weeks or more [5]; and the initial lack of Phase 3 trial data on single dose vaccine performance beyond 3 weeks for the Pfizer/BioNTech vaccine [1].

On short-term timescales, and in the absence of risk-structure or the potential for differential rates of waning immunity, if the efficacy from one dose is more than half the efficacy from two doses, then it is always preferable to prioritise vaccinating as many people as possible with one dose. Yet, given clear variation in the burden of severe outcomes caused by COVID-19, the prioritisation of dosing schedules merits quantitative evaluation; such analyses have been performed in a non-UK context [14–16].

In this paper, we study prioritisation of a one dose or two dose vaccination schedule given a fixed number of vaccine doses and with respect to a measure of maximising averted deaths. We performed this analysis in the context of the population of England and age-stratified risk mortality. We recognise that, whilst averted deaths are one important indicator to inform the SARS-CoV-2 vaccination programme, in reality there are multiple relevant indicators for vaccination as a major public health intervention (for example, averted hospital admissions, averted long-COVID cases and averted quality adjusted life years lost). We stress that the analysis we present can be readily applied to other contexts and refined using alternative assumptions and outcome criteria.

Using a simple algorithmic method, we sought generic insights into the benefits of prioritising either first or second vaccine doses according to two types of strategy for vaccine dose allocation: (i) giving as many people one dose or as many people two doses as permitted by the number of doses available (homogeneous strategy); (ii) adding flexibility to the allocation scheme by allowing for a given percentage of vaccine doses being used for first doses, with the remainder used for second doses (heterogeneous strategy). Throughout, we explored the sensitivity to the relative efficacy of the first vaccine dose (compared to the efficacy attained following two vaccine doses). We acknowledge that this is a simplified representation of a complex dynamic process, whereby new supplies of vaccine are being manufactured and distributed over time, where second dose efficacy may change depending on the inter-dose separation [5, 17] and where there can be an intrinsic feedback between vaccination and population-level incidence. Nevertheless, parsimonious model structures (such as the one used in this study) may be swiftly developed and applied, whereas models with additional complexities typically require longer development times, finer-resolution data to be reliably parameterised and can result in parameter inference becoming more computationally intensive [18]. Timely delivery of findings before a policy decision is taken can be worth more than using a more complex method and obtaining results afterwards, provided any methodological limitations are made clear [19]. In the discussion we expand on how the findings from these theoretical results need to be interpreted to apply to the situation facing England, the UK and other nations.

## Methods

### Data on age-dependent mortality risk

We base our analysis on the estimated age distribution of mortality due to COVID-19 in the UK, with a particular focus on the Joint Committee on Vaccination and Immunisation (JCVI) Phase 1 priority groups for vaccination [20]. The nine target groups within the first phase of the vaccination programme encompass care home residents and workers, health care workers, all those clinically extremely vulnerable (CEV) and with underlying health conditions (UHC), and all those aged 50 years and above.

Due to the absence of precise estimates for either the size of each priority group or the relative risk of COVID-19 mortality for individuals in each group (compared to the overall population average), we considered two different sets of assumptions around these two statistics (labelled ‘Age only’ and ‘Priority Group estimate’), with details of the two estimates provided in Table 1.

**Table 1:** Estimates of priority group population size and relative mortality risk. The age-only estimates were based on age-group data (mid-2019 estimates) for England [21] and the age distribution of mortality due to COVID-19 in the UK, during the period 1st September 2020 until 1st February 2021 (data provided by Public Health England, Personal Communication). We based the Priority Group estimate on age-structured mortality data in the second wave using priority group population estimates from DHSC [23]. As the Priority Group estimate included care home residents and staff, health care workers (HCW), the clinically extremely vulnerable (CEV) and those with underlying health conditions (UHC), the total number of individuals differed between the the Priority Group estimate and the age-only estimate. Note that the group 2 category “≥ 80 years & HCW” for the age group only estimate corresponded to just those aged 80 years and above, whereas for the Priority Group estimate “≥80 years & HCW” included healthcare workers and those aged 80 years or above. Similarly, for the group 4 category “70-74 years & CEV”, the age groups only estimate included only those aged 70-74 years, while for the Priority Group estimate “70-74 years & CEV” included both those aged 70-74 years old and the clinically extremely vulnerable. The relative risk of COVID-19 mortality for individuals in each group is measured relative to the overall population averaged mortality risk. We give population sizes to one decimal place and relative risk values to either one decimal place or two significant figures (dependent on which format provided greater precision).

| Priority Group, $p$ | Age groups only | | Priority Group estimate | |
| --- | --- | --- | --- | --- |
| | Size, $P_p$(millions) | Relative Risk, $RR_p$ | Size, $P_p$ (millions) | Relative Risk, $RR_p$ |
| 1. Care Homes | - | - | 0.7 | 17.0 |
| 2. $\geq 80$ years & HCW | 2.8 | 12.3 | 6.0 | 6.8 |
| 3. 75-79 years | 1.9 | 3.9 | 1.9 | 3.9 |
| 4. 70-74 years & CEV | 2.7 | 2.0 | 3.7 | 2.0 |
| 5. 65-69 years | 2.8 | 1.2 | 2.4 | 1.2 |
| 6. UHC | - | - | 6.2 | 1.0 |
| 7. 60-64 years | 3.0 | 0.77 | 1.5 | 0.77 |
| 8. 55-59 years | 3.6 | 0.43 | 2.0 | 0.43 |
| 9. 50-54 years | 3.9 | 0.25 | 2.3 | 0.25 |
| 0-49 years | 35.3 | 0.035 | 29.3 | 0.035 |

For the age-only model, estimation of risk was based solely on the age-distribution of mortality due to COVID-19 in England (using deaths within 28 days of a confirmed COVID-19 positive test, data from Public Health England (PHE)), during the period 1st September 2020 until 1st February 2021, compared to the underlying population pyramid for England using mid-2019 Office for National Statistics (ONS) population estimates (Fig. 1) [21]. It is evident that older age groups suffered the greatest mortality, with 60% of deaths due to COVID-19 in those over 80 years of age even though they only comprise 5% of the population.

**Fig. 1:**
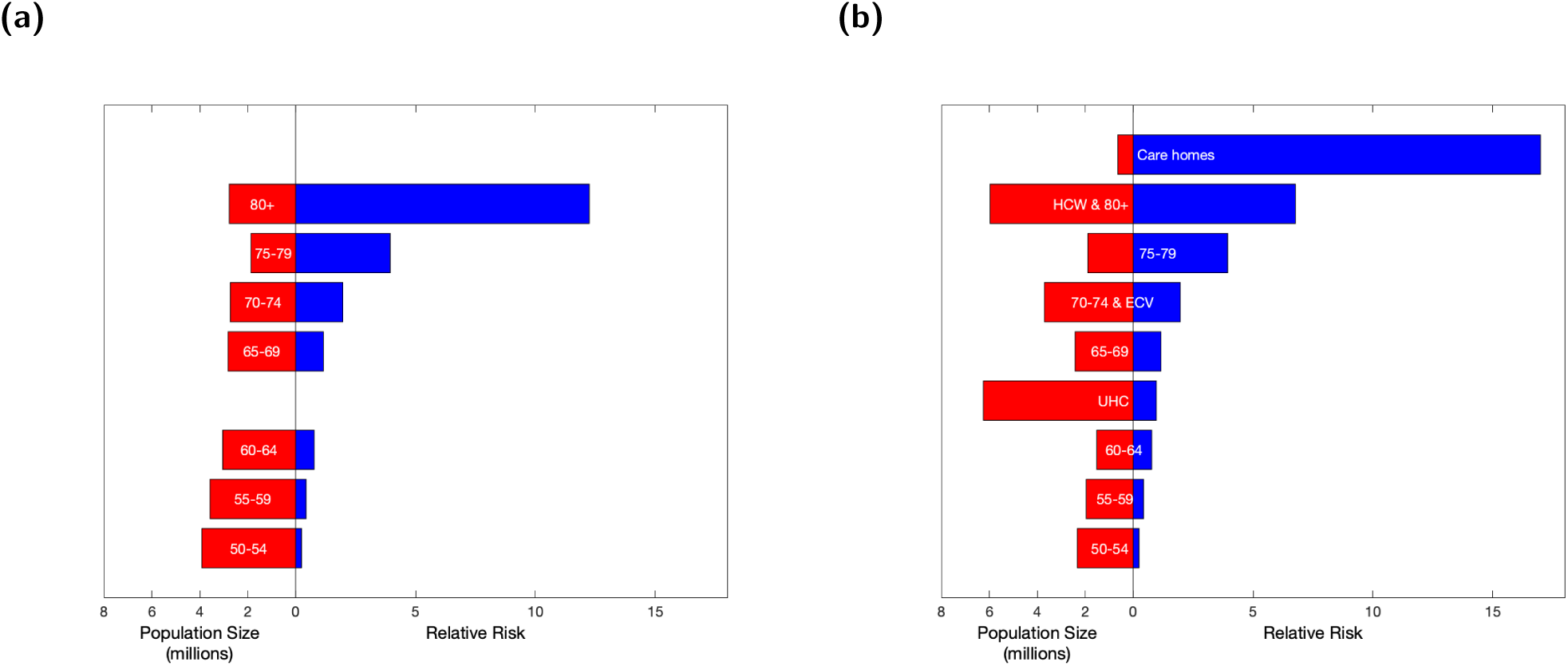
Graphical representation of the data in Table 1 showing the estimated population size (red) and the relative risk of mortality from COVID-19 (blue). **(a)** The assumptions for the age-structured estimates. **(b)** The assumptions for the Priority Group estimates. Vertical spacing of the two graphs is such that the groups of similar ages match.

For the Priority Group estimate, we amended the groups described for the age-only estimate to include the JCVI Phase 1 priority groups, assuming that this did not change the relative mortality risk of the age-groups (under 80 years old) previously calculated. Note that the Priority Group estimate including care home residents and staff, health care workers, the clinically extremely vulnerable and those with underlying health conditions meant the total number of individuals differed between the Priority Group estimate and the age-only estimate. The relative risk of care home residents and staff is based upon approximately 14 thousand care home deaths in the period since 7th August 2020 to the beginning of February 2021 [22], with the risk in the over-80s scaled to account for the greater risk of death within care homes. We assumed risks for those clinically extremely vulnerable to be equal to those aged 70-74, which also occupy priority group 4. We assumed risks for those with underlying health conditions (group 6) to lie equidistant between groups 5 and 7. Population estimates for these priority groups were provided by the Department of Health and Social Care (DHSC) [23].

### Vaccine assumptions

Using the population size data (*P*_*p*_) for each priority group, the associated relative risk of COVID-19 mortality (*RR*_*p*_) and estimates of vaccine efficacy following one or two doses (*VE*_1_ and *VE*_2_), we calculated the deaths averted given an assumed distribution of vaccines between the priority groups:

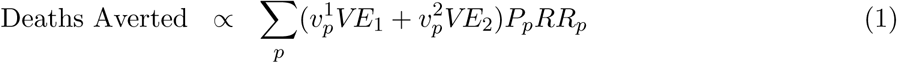

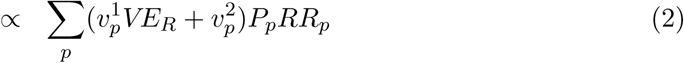

where 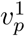 and 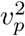 are the proportions of each priority group *p* that receive just one dose or two doses of the vaccine respectively. To further reduce the degrees of freedom of this calculation, it is sufficient to know the ratio of the vaccine efficacy from the first dose compared to the second *VE*_*R*_ = *VE*_1_*/VE*_2_, which we term relative efficacy of the first dose.

### Vaccine efficacy

At the original time of writing (in January 2021) the data on vaccine effectiveness in averting deaths due to SARS-CoV-2 infection following first and second dose with the vaccine was limited. We used the central estimates of vaccine efficacy against disease reported from clinical trial data to guide our range of relative efficacy: Pfizer/BioNTech (89% from first dose; 95% from two doses) [1]; Oxford/AstraZeneca (76% from first dose; 81% from two doses) [5]. This would imply that the relative efficacy of the first dose (*VE*_*R*_) is in the region of 93% for the Pfizer vaccine and the Oxford/AstraZeneca vaccine. In addition, there was data reported from the UK on mortality in those over 80 years old suggesting that the first dose of Pfizer vaccine reduced deaths by around 80%, which acted as a lower bound for the relative efficacy of the first dose against mortality [7].

This early data has now been superseded by more detailed analysis of the efficacy of the vaccines in the general population. For England, data on vaccine efficacy is calculated by PHE. Recent estimates of vaccine efficacy against COVID-19 mortality, as of July 2021, are 70-80% and 95-99% for one dose and two doses of the Pfizer/BioNTech vaccine, and 75-85% and 75-99% for one dose and two doses of the Oxford/AstraZeneca [24]. Taking the upper and lower bound of each range, this would give a relative efficacy of the first dose (*VE*_*R*_) of between 71% and 84% for the Pfizer vaccine, while for the AstraZeneca vaccine the estimate is between 76% and over 100% due to the greater uncertainty in the data. These estimates of efficacy against mortality due to COVID-19 have been generated for the Alpha (B.1.1.7) variant. For the Delta (B.1.617.2) variant, information on vaccine efficacy against mortality is still not available, but preliminary evidence suggests that the relative efficacy of the first dose may be lower.

### Strategies for vaccine dose allocation

We examined two types of strategy for dose allocation, which we describe as: (i) homogeneous strategy and (ii) heterogeneous strategy.

#### Homogeneous strategy

For a given number of available doses (*V*) and for a given relative efficacy from the first dose compared to the second (*VE*_*R*_), we first examined the question of whether to completely prioritise one dose or two doses of the vaccine. This essentially is a question of whether there is a greater number of expected deaths averted from giving as many people as possible one dose or two doses.

We compared the relative risks in the different age-groups (Table 1) and computed the relative number of deaths averted by the one dose and two dose strategies.

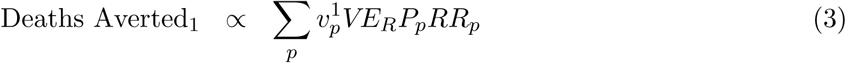

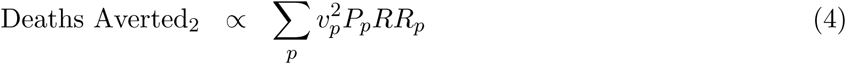

where there is a strict limit on the number of available doses: 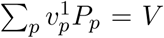 or 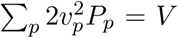. We note that this is a relative measure as predicting the scale of the future cases, hospitalisation and deaths is contingent on a number of policy decisions. In all scenarios we assumed 90% vaccine uptake (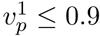 or 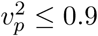), independent of age and priority group.

Given that the relative risk of COVID-19 mortality (*RR*_*p*_) decreases monotonically between risk groups, it is clear that the optimal deployment of either one of two doses must similarly decline monotonically (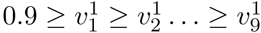 and similarly for the second dose 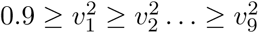). Moreover, it is always better to maximally vaccinate the higher-risk groups before preceding to lower-risk ones; therefore, solutions are generally of the form: 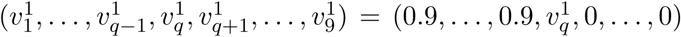 which corresponds to completely vaccinating groups 1 to *q -* 1, partially vaccinating group *q*, and not yet vaccinating the remaining lower-risk groups. This enables us to calculate the optimal deployment of vaccine across all priority groups without having to perform an exhaustive combinatorial search.

#### Heterogeneous strategy

We extended our initial analysis to consider a heterogeneous strategy. For a given number of doses, we sought the optimal deployment of a mixed scheme where some priority groups can be targeted for two doses while others receive one.

As an example, based on English population data, supposing we had 6.6 million doses of vaccine, these could either give all those aged 80 and above two doses, or it could give everyone aged 75 and above, and some of those in the 70-74 years age group, one dose. Again, our aim is to maximise the number of deaths averted, subject to the constraint on the total amount of vaccine available (*V*):

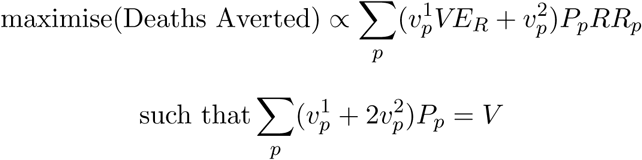

Again, due to the monotonicity on the relative risk of mortality, we can insist on a simple ordering of vaccination (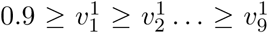 and 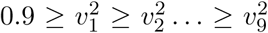); and again, we expect to maximally vaccinate higher risk groups before moving to lower ones. This essentially means we search over the number of vaccines allocated to second rather than first doses.

We studied the optimal allocation of vaccine for the two estimates of priority group size and relative risk (either based on age-structure only or using priority groups estimates), and for a range of relative efficacy of one dose compared to two doses (75% *≤ VE*_*R*_ *≤* 90%). We assumed vaccine uptake of 90% (to set the scale of vaccination in each priority group) and ignored the impact of transmission blocking (which is difficult to incorporate in this static model and is still not well quantified).

All computations and visualisations were performed in Matlab.

## Results

### Homogeneous strategy

For a given number of vaccine doses (*V*) and considering vaccine targeting towards age-group based priority groups, we considered when it is optimal to focus all vaccine resources on maximising the number of people receiving one dose or concentrate on ensuring that the most vulnerable groups get two doses.

When the number of vaccines available is insufficient to cover a specified age range or priority group, there is a choice between giving one dose to some proportion of the over 80’s or two doses to only half that number. In this situation, and ignoring the implications of generating long-term immunity, a one dose strategy would be favoured if 2*VE*_1_ *> VE*_2_ (*VE*_*R*_ *>* 0.5). For a larger available number of vaccine doses, we are faced with the dilemma between giving one dose to ages that are at slightly less risk or giving two doses to those that are most vulnerable. Using England once more as an example, supposing we had 5.5 million doses of vaccine, these could either be used to give all those aged 80 and above two doses, or could give everyone aged 75 and above, and some of those in the 70-74 years age group, one dose. To qualitatively assess this situation, we examined optimisation outcomes based on the two estimates for vaccination priority group population size and relative risk of mortality (Table 1).

For the age-only estimate of relative risk, the separation between prioritising first dose or second doses (Fig. 2(a)) was relatively smooth. For low numbers of available doses (*<* 2 million) and greater than 50% relative efficacy, the optimal policy is to prioritise one dose. For larger stockpiles of vaccine, the relative efficacy needs to be higher to prioritise giving one dose to as many people as possible. Within the plausible range of relative efficacy values (75% - 90%), we found a steady switch to prioritising the second dose as the amount of available vaccine increases from 4 million to 18 million doses.

**Fig. 2:**
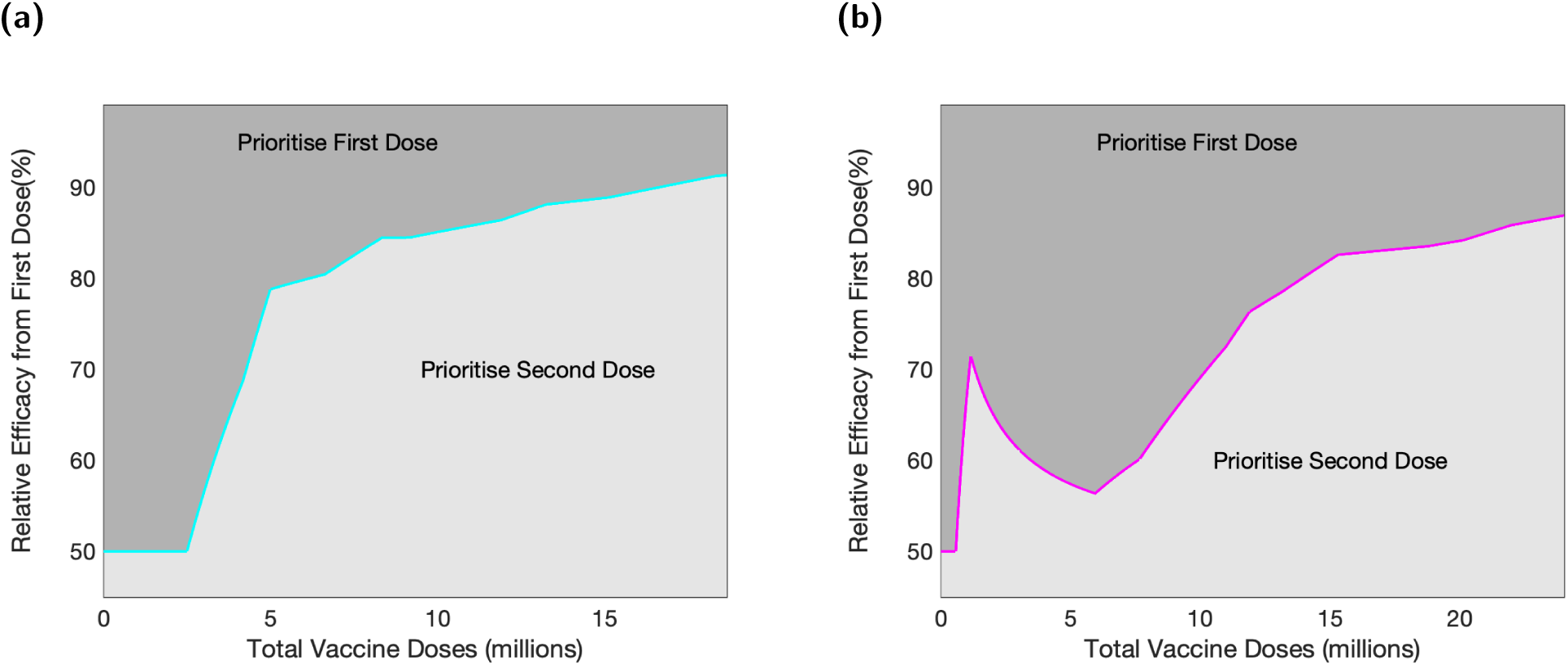
Optimisation of dosing strategy with respect to the number of vaccine doses and the relative efficacy of the first dose compared to the second dose. Panels correspond to outputs for two different estimates of vaccination priority group population size and relative risk of mortality (see Table 1 for further details). **(a)** Age groups only. **(b)** Priority Group estimate, which included specific groups for care homes and those with underlying health conditions. In all panels, and given a metric of maximising deaths averted, dark shaded regions correspond to parameter sets where it was determined optimal to prioritise first doses, with light shaded regions corresponding to parameter sets where two dose vaccination was optimal. The maximal number of doses considered corresponds to being able to give all individuals in the priority groups one dose, assuming 90% uptake.

For the Priority Group estimate (Fig. 2(b)), we observed a broadly similar pattern; however, the very high relative risk associated with care home residents and workers (priority group 1) means that, for a low number of doses and a low relative efficacy, it can be optimal to prioritise giving two doses to the care home group. With this estimated set of relative risks, there was also an even stronger effect (compared to the age-only estimate) of high relative first dose efficacy, leading to a wider parameter space where the first dose was prioritised.

### Heterogeneous strategy

We next considered strategies where a given proportion of the available vaccine are used for first doses and the remainder for second doses. We performed this assessment under an assumption of maximising the number of deaths averted and a vaccine uptake of 90%.

Given a relative efficacy for the first dose of below 50%, the optimal strategy is to use half of the available vaccine for second doses, such that everyone prioritised for vaccination receives two doses (Fig. 3). Above this threshold of 50% relative efficacy from the first dose, the pattern of doses reserved for second doses approximately follows the same pattern as the homogeneous strategy (cyan and pink lines in Fig. 3 are the same as in Fig. 2). We found a smaller region of parameter space where the optimal strategy is to only give one dose (dark blue, and only for a low number of doses or very high levels of relative efficacy of the first dose). The distinct banding observed is due to the switch between different priority groups as the amount of available vaccine increases. For the Priority Group estimate, as with the homogeneous strategy, a distinct structure was visible in the results: a two dose strategy (focused on care homes) was optimal at around 2 million doses and for a relative first-dose efficacy of up to 70% (Fig. 3(b)).

**Fig. 3:**
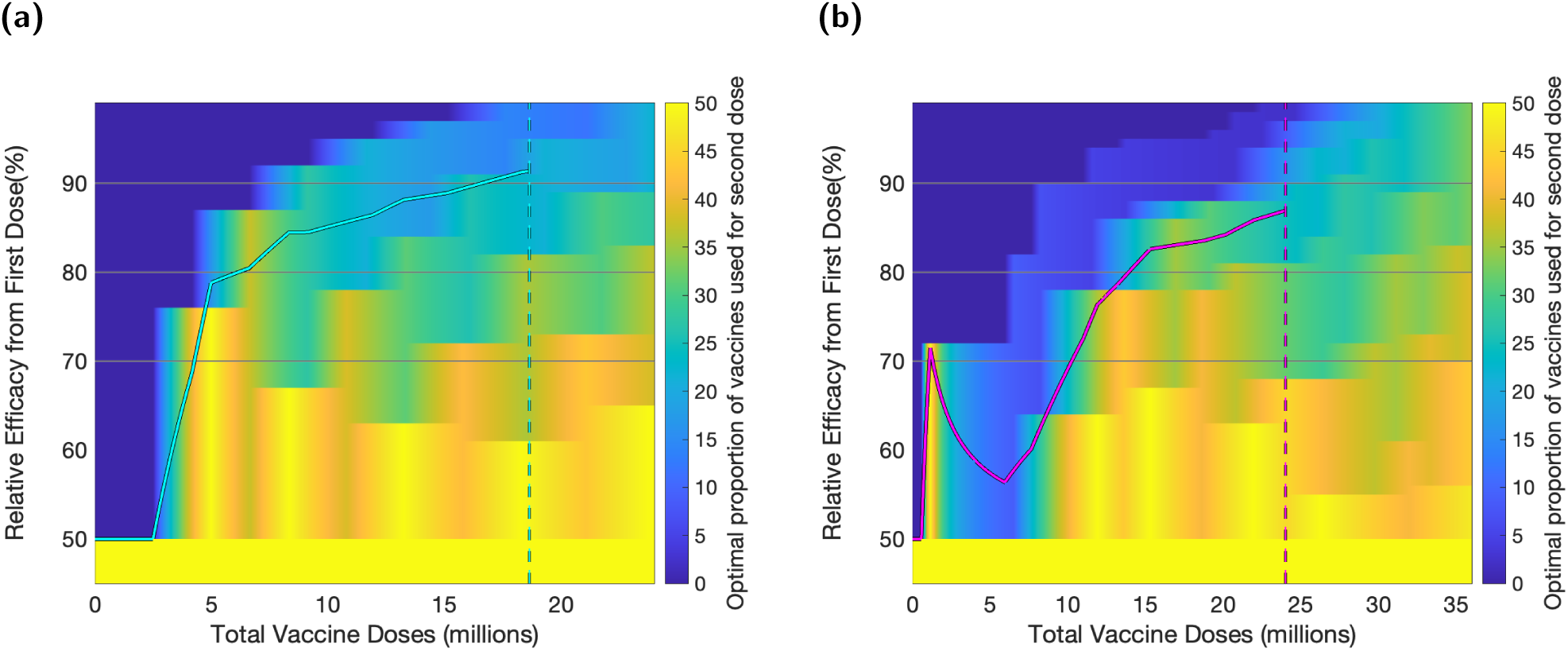
Optimal distribution of a given number of vaccine doses between first and second dose. Regions of parameter space in which most doses should be prioritised toward first dose are coloured in dark blue, with gradation to yellow for an increasing proportion of doses being used as second doses. The solid lines show the boundary between the parameter regions associated with homogeneous strategies shown in Fig. 2. **(a)** Age groups only; **(b)** Priority Group estimate. Relative efficacy of 70%, 80% and 90% are highlighted (horizontal lines) for comparison with later plots. Vertical dashed lines shows the number of vaccine doses required to give all of the nine priority groups (in Phase 1 of the vaccination programme) one dose, assuming 90% uptake.

For a given ratio of first and second doses, the associated distribution of vaccine between the priority groups can again be calculated due to the monotonicity of the relative risk. We show the optimal distribution for a distinct set of relative efficacies from the first dose (*VE*_*R*_ = 70%, 80%, 90%) and for a specified number of doses (4, 8, 12, 16, 20 and 24 million) (Fig. 4). We show as stacked bars the number of first (left) and second (right) doses given to each priority group for both the simple age-structured estimates of risk and for the full Priority Group estimates.

**Fig. 4:**
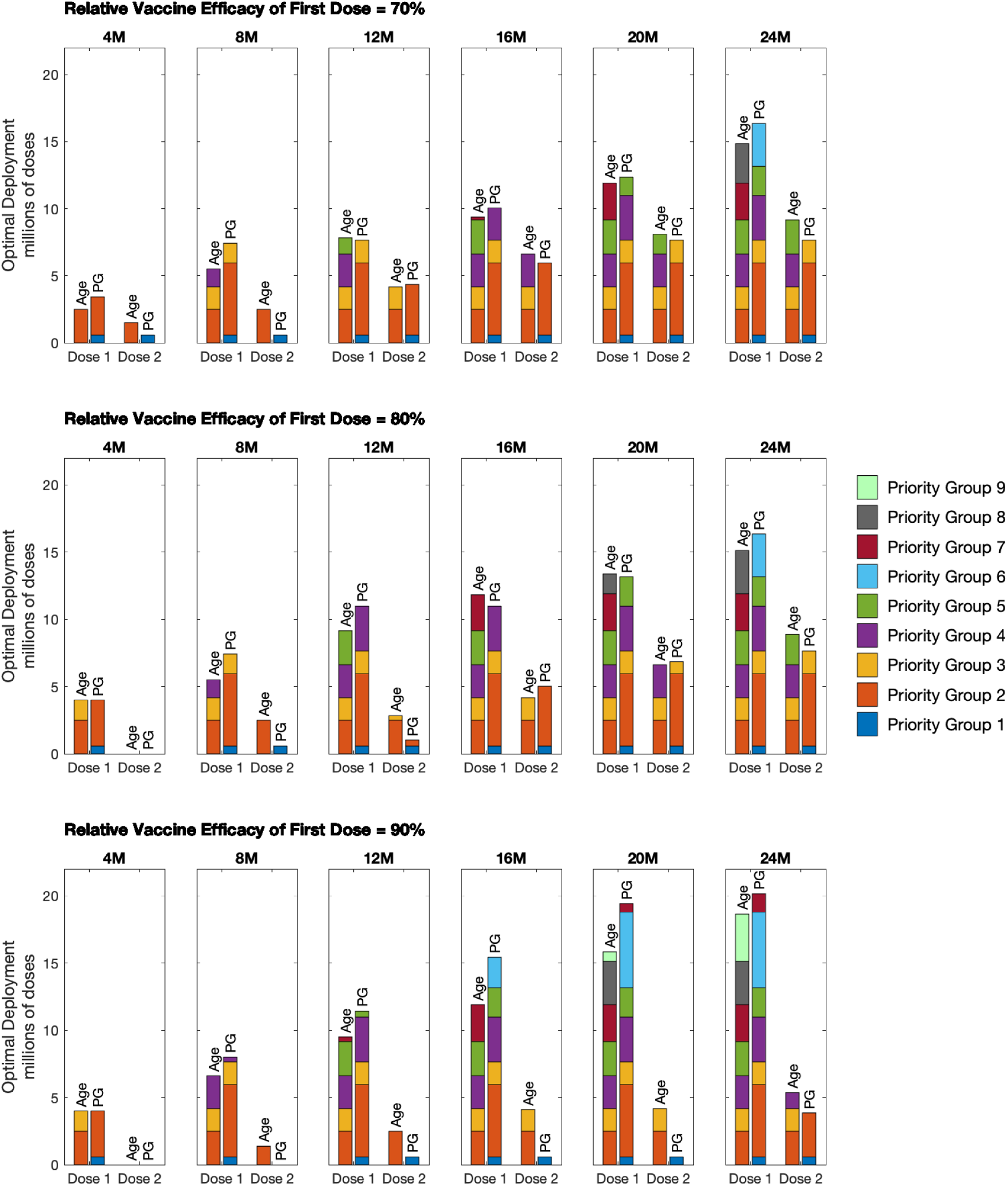
Optimal deployment of vaccine for the two different priority group estimates and a selection of vaccine efficacies. We considered a relative efficacy of first dose to second dose of: **(a)** 70%; **(b)** 80%; **(c)** 90%. For each pair of bar plots, the left bar corresponds to Age Group estimates and the right bar corresponds to Priority Groups.

At 70% relative efficacy, there was a strong tendency to offer second doses shortly after the first. Thus at 4 million doses, the optimal strategy was to begin offering second doses to either the oldest age-group or priority group 1. For higher levels of vaccine availability (e.g. 24 million doses), although the distribution of second doses lags behind the first, we consistently predict at least 50% of the groups receiving two doses of vaccine is optimal.

When relative efficacy is higher (80% or 90%) there is more of a delay before it becomes optimal to begin second vaccinations. At 4 million doses, the optimal strategy became focused on delivering single doses only; with second doses being introduced more gradually. At the most extreme parameters investigated (90% relative vaccine efficacy and full priority group estimates), even at 20 million doses, the only group to have received their second dose was priority group 1 (care home residents and staff).

Although we generated these figures by simply considering the optimal use of a fixed pool of available vaccine - with no reference to how lower amounts of vaccine have been used - it is still possible to read the graphs as a chronological sequence, due to the monotonicity of the relative risk. As such, for any given relative efficacy, the first *V* doses of vaccine are always distributed in the same manner (Fig. 5). An alternative way to view the same information is to consider at what point in the delivery programme it becomes optimal to give first and second doses to each of the priority groups. For the relative risk of mortality estimated for the full priority groups, this visualisation clarifies that at high relative efficacy from the first dose of vaccine (90%) the optimal distribution of vaccine is substantially weighted towards early prioritisation of first doses with a substantial delay until the second dose is offered. For completion of the first four priority groups (everyone over 70, health care workers, care home staff and residents and those that are clinically extremely vulnerable) we estimate that the optimal delay between finishing the first doses and finishing the second doses is: 12.83 million doses (for a relative efficacy of 70%); 19.58 million doses (for a relative efficacy of 80%) and 24.01 million doses (for a relative efficacy of 90%) - which is between 6 and 12 weeks if delivery is maintained at 2 million doses a week.

**Fig. 5:**
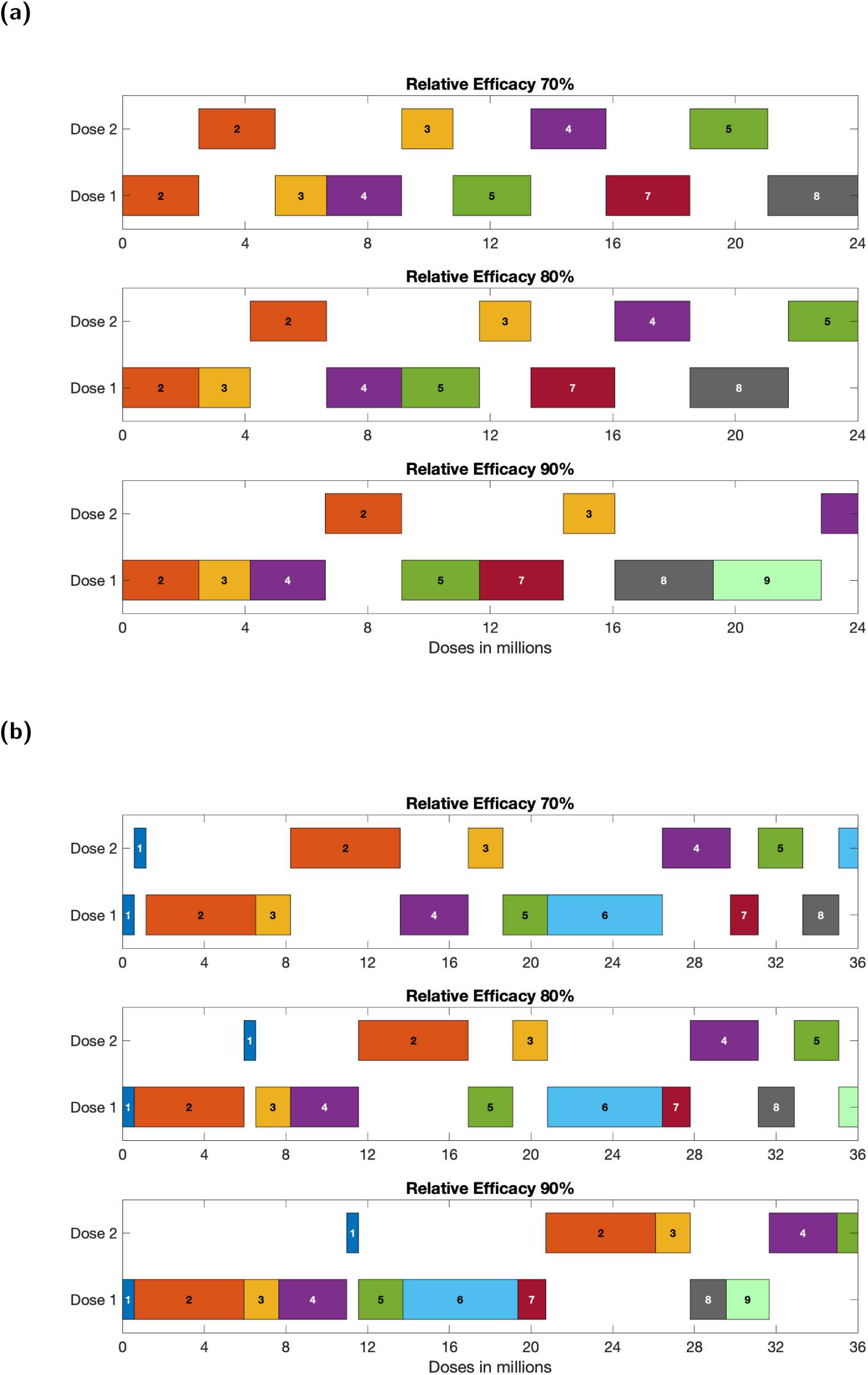
Dose allocation schedule, dependent upon relative first dose efficacy. **(a)** Age only estimate of relative risk. **(b)** Priority Group estimate of relative risk. At any point in the delivery schedule each pane shows the optimal groups that should be prioritised for vaccination. We show a smaller range of total doses for (a) compared to (b), as the total number of individuals over 50 years old is smaller than the number in priority groups 1-9.

## Discussion

Here we have developed a simple algorithmic method that can optimise the distribution of a fixed number of vaccine doses, allowing us to maximise averted deaths. The JCVI Phase 1 priority groups have been defined such that early groups have a higher risk of mortality than lower ones [20, 25]. There is hence a natural ordering in which we would wish to vaccine priority group 1 before moving to priority group 2. The more challenging question that we address here is whether it is better to give high-risk groups their second dose of vaccine before giving lower-risk groups (in the priority order) their first dose. In the context of UK vaccination policy, the key question was around the delay between the first and second dose, with a longer delay allowing more individuals to be given some level of protection in the short-term. For the Oxford/AstraZeneca vaccine there is also compelling evidence that a delay of 12 weeks or more provides greater second dose vaccine efficacy [5], strengthening the case for an early prioritisation of first doses; this has since been enhanced by studies of the Pfizer vaccine suggesting a longer delay may again offer better protection [17].

In countries where the total supply of vaccine is more limited, similar calculations could inform whether a strategy that attempted to maximise coverage by only giving a single dose would be of benefit - although, in this scenario, far more information would be required on the long-term protection offered by a single dose.

The key parameter in our model is the relative efficacy provided by the first dose of vaccine compared to the level of protection offered by two doses. Here, we have focused on COVID-19 mortality using the relative risk of infection followed by death for each of the nine JCVI priority groups, and hence we are most interested in efficacy against death. Unfortunately, efficacy against death is extremely difficult to measure from Phase 3 trials (no-one taking part in the Pfizer/BioNTech trials, in either the control or vaccine arm, died with COVID-19 [1], with one COVID-19-related death in one participant in the control group of Oxford/AstraZeneca trials [5]), and so we need to rely on data from the large-scale national programmes. Early data from the UK on those over 80 years of age (and therefore amongst the first to receive the vaccine) suggested that a first dose of the Pfizer/BioNTech vaccine generates a vaccine efficacy against symptomatic infection of 70% (95% CI 59-78%) after four weeks and an additional 51% (95% CI 37-62%) lower risk of death if infected, giving a combined efficacy against death of 85% [7]. This is therefore a lower-bound on our required relative efficacy. These early estimates have since been superseded with estimates available for both Pfizer/BioNTech and Oxford/AstraZeneca vaccines after one and two doses for the Alpha variant [24]; however even in this well studied example their remains considerable uncertainty in the relative efficacy of the first dose. For other variants of concern the uncertainty in vaccine efficacy is even greater.

We predict that, for relatively high protection from the first dose (compared to the efficacy derived from two doses), a substantial number of first doses should be administered before attention switches to giving second doses (Fig. 5). We expect these simple trade-offs to occur at any point in the vaccination program where logistical constrains (vaccine supply or number of trained vaccinaters) limits uptake. As such, under these circumstances, early vaccine roll-out ought to be targeted towards giving as many people one dose as possible, until the switch-point is reached. Our results agree with findings from earlier modelling work (applied in a non-UK context) that found, when a single dose retains the majority of the effectiveness against disease of two doses, immunising as many individuals as possible with a single-dose regimen may achieve a greater reduction in disease from COVID-19 than a two-dose regimen in a smaller population [14–16].

While this modelling provides important generic insights into the benefits of first and second doses, there are a number of elements that are absent from this simple analysis. Most notably, the vaccination programme is a dynamic process in which different amounts of vaccine are available at different points in time; therefore, while it is possible to read Fig. 5 as a chronology, it does not take into account the necessary biological restrictions on the appropriate separation between doses. Our model computes the protection derived from a specified amount of vaccine doses being instantaneously administered amongst the population. This lack of a dynamic perspective means that we cannot address questions that relate to the precise timing of vaccination. In particular, very long delays between doses may have implications for both short-and long-term immunity; similarly the model cannot directly capture the delay between vaccination and the development of immunity. Subsequent modelling studies have since demonstrated the importance of quantifying the characteristics and durability of vaccine-induced protection after the first vaccine dose in order to determine the optimal time interval between the two vaccine doses [26]. In addition, our model also assumes that priority groups are completed in order of greatest risk - whereas in practice, and for a number of practical reasons, the delivery schedule is blurred, often vaccinating groups that are most easy to reach. We expect a schedule that mixes priority groups to lessen the advantage of prioritising the delivery of first doses compared to re-vaccinating with second doses.

Our determination of dose allocation was based on averting deaths, with no regard for hospital admissions (and therefore pressure on the health services), the implications of long-COVID nor any form of life-years lost or quality adjusted life year assessment. The prioritisation of first doses compared to second doses, for a given relative efficacy, may differ under an alternative metric or collection of measures (as found in the study by Matrajt *et al*. [16], who determined that the optimal allocation strategy with one and two doses of vaccine was different when minimising one of five distinct metrics of disease and healthcare burden under various degrees of viral transmission) and is a topic for follow-up work. For an objective of minimising COVID-19 morbidity, then a similar pattern of prioritisation is expected; measures of COVID-19 morbidity are consistently higher in the elderly and vulnerable risk groups such that these groups should be prioritised to receive their first dose of vaccine as early as possible. However, many measures of morbidity risk (such as hospital admissions) are not as skewed towards older age-groups as the mortality risk we have used throughout this document, although they are still highly age-dependent. A similar change to age-dependent factors would occur if we considered the objective of minimising life-years lost, as mortality in older age groups results in fewer life-years lost than mortality in younger age groups, although analysis suggests that life-years lost to COVID-19 is still an increasing function of age. For these less skewed measures, there is less incentive to rapidly give older individuals a second dose as the relative risk in older individuals compared to younger individuals is not as high. By this reasoning, we expect an objective of minimising life-years lost or COVID-19 morbidity to increase the prioritisation of first doses over second doses, with second doses in the older individuals not generating the greatest benefit until a larger proportion of the population have been given first doses.

In terms of the demography and empirical data on mortality risk due to COVID-19, our analysis has been carried out using data corresponding to the population of England. Thus, these findings will not necessarily directly translate to other settings, in particular where the population structure and mortality are vastly different. Finally, our static modelling framework does not account for the transmission dynamics of infection; the fact that individuals have been immunised does not change the risk to the remaining population and hence we do not capture the structured reduction in risk that can occur, although general declines (or increases) in risk that apply equally to the entire population do not affect our results. As such, our approach does not address questions such as herd immunity nor how vaccination will impact the long-term trajectory of the outbreak [27]. Instead, the methods are tailored toward the early stages of a vaccination programme where the aim is to reduce mortality as rapidly as possible.

In summary, given the strong evidence that a single dose is highly effective, our model results would indicate that early prioritisation of one dose (compared to re-vaccinating with a second doses) averts the greater number of deaths. The precise timing of first and second doses is contingent on the speed of the delivery programme, with more rapid delivery favouring early deployment of second doses. The policy adopted in the UK was dependent upon a number of practical considerations - not least the greater second dose efficacy of the Oxford/AstraZeneca vaccine after a 12-week delay [5], and the need for a simple, consistent message across all priority groups and vaccines. However, this work clearly shows that, given particular combinations of demographic and vaccine attributes, a strategy of prioritising first doses can have substantial public health benefits.

## Data Availability

The data used to conduct this study are provided in the main manuscript. Code is available at https://github.com/EdMHill/fixed_num_vaccine_doses_one_vs_two_dose_prioritisation.

## Author contributions

**Edward M. Hill:** Methodology, Software, Validation, Writing - Original Draft, Writing - Review & Editing.

**Matt J. Keeling:** Conceptualisation, Methodology, Software, Formal analysis, Visualisation, Writing - Original Draft, Writing - Review & Editing.

**Acknowledgements**

Thanks to Ian Hall and Department of Health and Social Care for assisting with provision of priority group population estimates.

## Data and code accessibility

The data used to conduct this study are provided in the main manuscript. Code is available at https://github.com/EdMHill/fixed num vaccine doses one vs two dose prioritisation.

## References

[1] Polack FP, Thomas SJ, Kitchin N, Absalon J, Gurtman A, et al. Safety and Efficacy of the BNT162b2 mRNA Covid-19 Vaccine. N. Engl. J. Med. 383(27):2603–2615 (2020). doi:10.1056/NEJMoa2034577.

[2] Ramasamy MN, Minassian AM, Ewer KJ, Flaxman AL, Folegatti PM, et al. Safety and immunogenicity of ChAdOx1 nCoV-19 vaccine administered in a prime-boost regimen in young and old adults (COV002): a single-blind, randomised, controlled, phase 2/3 trial. Lancet 396(10267):1979–1993 (2020). doi:10.1016/S0140-6736(20)32466-1.

[3] Baden LR, El Sahly HM, Essink B, Kotloff K, Frey S, et al. Efficacy and Safety of the mRNA-1273 SARS-CoV-2 Vaccine. N. Engl. J. Med. 384(5):403–416 (2021). doi:10.1056/NEJMoa2035389.

[4] Voysey M, Clemens SAC, Madhi SA, Weckx LY, Folegatti PM, et al. Safety and efficacy of the ChAdOx1 nCoV-19 vaccine (AZD1222) against SARS-CoV-2: an interim analysis of four randomised controlled trials in Brazil, South Africa, and the UK. Lancet 397(10269):99–111 (2021). doi:10.1016/S0140-6736(20)32661-1.

[5] Voysey M, Costa Clemens SA, Madhi SA, Weckx LY, Folegatti PM, et al. Single-dose administration and the influence of the timing of the booster dose on immunogenicity and efficacy of ChAdOx1 nCoV-19 (AZD1222) vaccine: a pooled analysis of four randomised trials. Lancet 397(10277):881–891 (2021). doi:10.1016/S0140-6736(21)00432-3.

[6] Rossman H, Shilo S, Meir T, Gorfine M, Shalit U, et al. COVID-19 dynamics after a national immunization program in Israel. Nat. Med. 27(6):1055–1061 (2021). doi:10.1038/s41591-021-01337-2.

[7] Lopez Bernal J, Andrews N, Gower C, Robertson C, Stowe J, et al. Effectiveness of the Pfizer-BioNTech and Oxford-AstraZeneca vaccines on covid-19 related symptoms, hospital admissions, and mortality in older adults in England: test negative case-control study. BMJ 373:n1088 (2021). doi:10.1136/bmj.n1088.

[8] Medicines and Healthcare products Regulatory Agency. Regulatory approval of Pfizer/BioNTech vaccine for COVID-19 (2021). URL https://www.gov.uk/government/publications/regulatory-approval-of-pfizer-biontech-vaccine-for-covid-19. [Online] (Accessed: 03 August 2021).

[9] Medicines and Healthcare products Regulatory Agency. Regulatory approval of Vaxzevria (previously COVID-19 Vaccine AstraZeneca) (2021). URL https://www.gov.uk/government/publications/regulatory-approval-of-covid-19-vaccine-astrazeneca. [Online] (Accessed: 03 August 2021).

[10] Public Health England. Annex A: Report to JCVI on estimated efficacy of a single dose of Pfizer BioNTech (BNT162b2 mRNA) vaccine and of a single dose of ChAdOx1 vaccine (AZD1222) (2021). URL https://assets.publishing.service.gov.uk/government/uploads/system/uploads/attachmentdata/file/949505/annex-a-phe-report-to-jcvi-on-estimated-efficacy-of-single-vaccine-dose.pdf. [Online] (Accessed: 03 August 2021).

[11] Office for National Statistics. Coronavirus (COVID-19) Infection Survey, UK: 8 January 2021 (2021). URL https://www.ons.gov.uk/peoplepopulationandcommunity/healthandsocialcare/conditionsanddiseases/bulletins/coronaviruscovid19infectionsurveypilot/8january2021. [Online] (Accessed: 03 August 2021).

[12] UK Government. Coronavirus (COVID-19) in the UK dashboard: Healthcare (2021). URL https://coronavirus.data.gov.uk/details/healthcare. [Online] (Accessed: 03 August 2021).

[13] UK Government. Coronavirus (COVID-19) in the UK dashboard: Deaths (2021). URL https://coronavirus.data.gov.uk/details/deaths. [Online] (Accessed: 03 August 2021).

[14] Paltiel AD, Zheng A, Schwartz JL. Speed Versus Efficacy: Quantifying Potential Tradeoffs in COVID-19 Vaccine Deployment. Ann. Intern. Med. 174(4):568–570 (2021). doi: 10.7326/M20-7866.

[15] Tuite AR, Fisman DN, Zhu L, Salomon JA. Alternative Dose Allocation Strategies to Increase Benefits From Constrained COVID-19 Vaccine Supply. Ann. Intern. Med. 174(4):570–572 (2021). doi:10.7326/M20-8137.

[16] Matrajt L, Eaton J, Leung T, Dimitrov D, Schiffer JT, et al. Optimizing vaccine allocation for COVID-19 vaccines: critical role of single-dose vaccination. medRxiv page 2020.12.31.20249099 (2021). doi:10.1101/2020.12.31.20249099.

[17] Parry H, Bruton R, Stephens C, Brown K, Amirthalingam G, et al. Extended interval BNT162b2 vaccination enhances peak antibody generation in older people. medRxiv page 2021.05.15.21257017 (2021). doi:10.1101/2021.05.15.21257017.

[18] Funk S, King AA. Choices and trade-offs in inference with infectious disease models. Epidemics 30:100383 (2020). doi:10.1016/j.epidem.2019.100383.

[19] Whitty CJM. What makes an academic paper useful for health policy? BMC Med. 13:301 (2015). doi:10.1186/s12916-015-0544-8.

[20] Department of Health & Social Care. Priority groups for coronavirus (COVID-19) vaccination: advice from the JCVI, 30 December 2020 (2021). URL https://www.gov.uk/government/publications/priority-groups-for-coronavirus-covid-19-vaccination-advice-from-the-jcvi-30-december-2020/joint-committee-on-vaccination-and-immunisation-advice-on-priority-groups-for-covid-19-vaccination-30-dec [Online] (Accessed: 03 August 2021).

[21] Office for National Statistics. Dataset: Estimates of the population for the UK, England and Wales, Scotland and Northern Ireland (2021). URL https://www.ons.gov.uk/peoplepopulationandcommunity/populationandmigration/populationestimates/datasets/populationestimatesforukenglandandwalesscotlandandnorthernireland. [Online] (Accessed: 03 August 2021).

[22] Office for National Statistics. Dataset: Number of deaths in care homes notified to the Care Quality Commission, England (2021). URL https://www.ons.gov.uk/peoplepopulationandcommunity/birthsdeathsandmarriages/deaths/datasets/numberofdeathsincarehomesnotifiedtothecarequalitycommissionengland. [Online] (Accessed: 03 August 2021).

[23] Department of Health and Social Care. UK COVID-19 vaccines delivery plan (2021). URL https://assets.publishing.service.gov.uk/government/uploads/system/uploads/attachmentdata/file/951928/uk-covid-19-vaccines-delivery-plan-final.pdf. [Online] (Accessed: 03 August 2021).

[24] Public Health England. COVID-19 vaccine surveillance report: 1 July 2021 (week 26) (2021). URL https://assets.publishing.service.gov.uk/government/uploads/system/uploads/attachmentdata/file/998411/Vaccine_surveillance_report_-_week26.pdf. [Online] (Accessed: 03 August 2021).

[25] Department of Health & Social Care. Annex A: COVID-19 vaccine and health inequalities: considerations for prioritisation and implementation (2021). URL https://www.gov.uk/government/publications/priority-groups-for-coronavirus-covid-19-vaccination-advice-from-the-jcvi-30-december-2020/annex-a-covid-19-vaccine-and-health-inequalities-considerations-for-prioritisation-and-implementation. [Online] (Accessed: 03 August 2021).

[26] Moghadas SM, Vilches TN, Zhang K, Nourbakhsh S, Sah P, et al. Evaluation of COVID-19 vaccination strategies with a delayed second dose. PLOS Biol. 19(4):e3001211 (2021). doi: 10.1371/journal.pbio.3001211.

[27] Moore S, Hill EM, Tildesley MJ, Dyson L, Keeling MJ. Vaccination and non-pharmaceutical interventions for COVID-19: a mathematical modelling study. Lancet Infect. Dis. 21(6):793–802 (2021). doi:10.1016/S1473-3099(21)00143-2.

